# Identifying Proteomic Prognostic Markers for Alzheimer’s Disease with Survival Machine Learning: the Framingham Heart Study

**DOI:** 10.1101/2024.09.21.24314123

**Authors:** Yuanming Leng, Huitong Ding, Ting Fang Alvin Ang, Rhoda Au, P. Murali Doraiswamy, Chunyu Liu

**Affiliations:** Department of Biostatistics, Boston University School of Public Health, Boston, MA, 02118, USA; Department of Anatomy and Neurobiology, Boston University Chobanian & Avedisian School of Medicine, Boston, MA, 02118, USA; Framingham Heart Study, Boston University Chobanian & Avedisian School of Medicine, Boston, MA, 02118, USA; Slone Epidemiology Center, Boston University Chobanian & Avedisian School of Medicine, Boston, MA, 02118, USA; Departments of Neurology and Medicine, Boston University Chobanian & Avedisian School of Medicine, Boston, MA, 02118, USA; Department of Epidemiology, Boston University School of Public Health, Boston, MA, 02118, USA; Department of Psychiatry, Neurocognitive Disorders Program, Duke University School of Medicine, Durham, NC, 27710, USA

**Keywords:** Alzheimer’s disease, Proteomics, Prognostic markers, Risk, Survival machine learning

## Abstract

**Background:** Protein abundance levels, sensitive to both physiological changes and external interventions, are useful for assessing the Alzheimer’s disease (AD) risk and treatment efficacy. However, identifying proteomic prognostic markers for AD is challenging by their high dimensionality and inherent correlations.

**Methods:** Our study analyzed 1128 plasma proteins, measured by the SOMAscan platform, from 858 participants 55 years and older (mean age 63 years, 52.9% women) of the Framingham Heart Study (FHS) Offspring cohort. We conducted regression analysis and machine learning models, including LASSO-based Cox proportional hazard regression model (LASSO) and generalized boosted regression model (GBM), to identify protein prognostic markers. These markers were used to construct a weighted proteomic composite score, the AD prediction performance of which was assessed using time-dependent area under the curve (AUC). The association between the composite score and memory domain was examined in 339 (of the 858) participants with available memory scores, and in an independent group of 430 participants younger than 55 years (mean age 46, 56.7% women).

**Results:** Over a mean follow-up of 20 years, 132 (15.4%) participants developed AD. After adjusting for baseline age, sex, education, and APOE ε4+ status, regression models identified 309 proteins (*P* ≤ 0.2). After applying machine learning methods, nine of these proteins were selected to develop a composite score. This score improved AD prediction beyond the factors of age, sex, education, and APOE ε4+ status across 15 to 25 years of follow-up, achieving its peak AUC of 0.84 in the LASSO model at the 22-year follow-up. It also showed a consistent negative association with memory scores in 339 participants (beta = -0.061, *P* = 0.046), 430 independent participants (beta = -0.060, *P* = 0.018), and the pooled 769 samples (beta = -0.058, *P* = 0.003).

**Conclusion:** These findings highlight the utility of proteomic markers in improving AD prediction and emphasize the complex pathology of AD. The composite score may aid early AD detection and efficacy monitoring, warranting further validation in diverse populations.

## Introduction

Alzheimer’s disease (AD) is a progressive neurodegenerative disorder that gradually impairs cognitive functions such as memory and reasoning abilities[1]. This disease significantly affects patients’ ability to perform daily tasks independently [2] and imposes a considerable burden on caregivers and healthcare systems [3]. With the global population aging, the incidence of AD is rising, posing a growing threat to public health and necessitating preventive strategies and effective treatment [4]. The complex nature of AD, especially its prolonged asymptomatic phase, presents challenges for early detection but also opportunities to develop interventions aimed at modifying the disease’s trajectory for secondary prevention [5, 6]. Therefore, identifying AD prognostic markers is crucial due to the disease’s insidious onset and progression and the lack of effective treatments for AD [7].

Plasma proteomic markers are sensitive to both internal physiological changes and external interventions[8], making them excellent candidates for tracking AD progression and response to treatment. Mang studies have highlighted significant associations between specific proteins and the risk of AD, along with associations between changes in protein levels and structural brain alterations over time[9–13]. Research has also identified a relationship between proteomic markers and amyloid burden, suggesting that plasma protein testing could be used to assess brain amyloid deposition[14]. Despite these advancements, the challenge of identifying proteomic prognostic markers remains, largely due to their high dimensionality and strong correlations among proteins. Moreover, there is a significant gap concerning the long-term predictive capacity of these proteomic markers for AD. This gap highlights the need for further research to evaluate how these markers perform in predicting AD incidence at specific future time points, potentially improving early detection and timely intervention strategies for AD.

Survival machine learning is particularly effective at addressing these challenges[15]. These methods are able to account for higher-order interactions and nonlinear relationships, which are crucial for selecting features based on variable importance[16]. Furthermore, survival machine learning models can incorporate time-to-event data and consider censored data. Therefore, we conducted association analysis and applied machine learning models to identify proteomic prognostic markers and constructed a weighted proteomic composite score in a community-based cohort. We aim to identify proteomic biomarkers and construct a composite score to enhance AD prediction across various follow-up periods.

## Methods

### Study Population

Initiated in 1948, the Framingham Heart Study (FHS) was a prospective cohort study based in a community setting[17]. In 1971, the study expanded to include the FHS Offspring cohort, comprising the children of the original participants and the spouses of these children [18]. Since the first Offspring exam cycle between 1971 and 1975, participants have undergone 10 health examinations approximately every four to six years[19]. This study included 1,913 individuals from the Offspring cohort who participated in the fifth examination cycle between 1991 and 1995, during which their blood was collected for proteomics profiling assessments. Participants were excluded if they had prevalent AD, incident non-AD dementia (n = 62), lacked education (n = 59) and APOE (n= 65) information. In analyses with machine learning, models tend to bias towards the majority class, which can lead to inflated performance metrics during training[20]. To address this imbalance and minimize potential model bias, we excluded participants who were 55 years or younger at baseline (n = 869), leaving 858 participants to identify protein markers to predict AD (**Figure 1**). This helped balance the age differences between cases and controls without significantly reducing the number of incident AD cases. For the association analysis of the proteomic composite score and memory domain, 339 of 858 participants with neuropsychological (NP) test measures were included (**Figure 1**). Additionally, to validate the early detection capabilities of the proteomic composite score for AD, we incorporated a separate, younger group of 430 independent participants into this analysis (**Figure 1**). All participants provided their written consent for genetic studies. The study protocol received approval from the Institutional Review Boards at Boston University Medical Center, Massachusetts General Hospital, and Beth Israel Deaconess Medical Center. The study adhered strictly to regulations and guidelines to ensure compliance.

**Figure 1.**
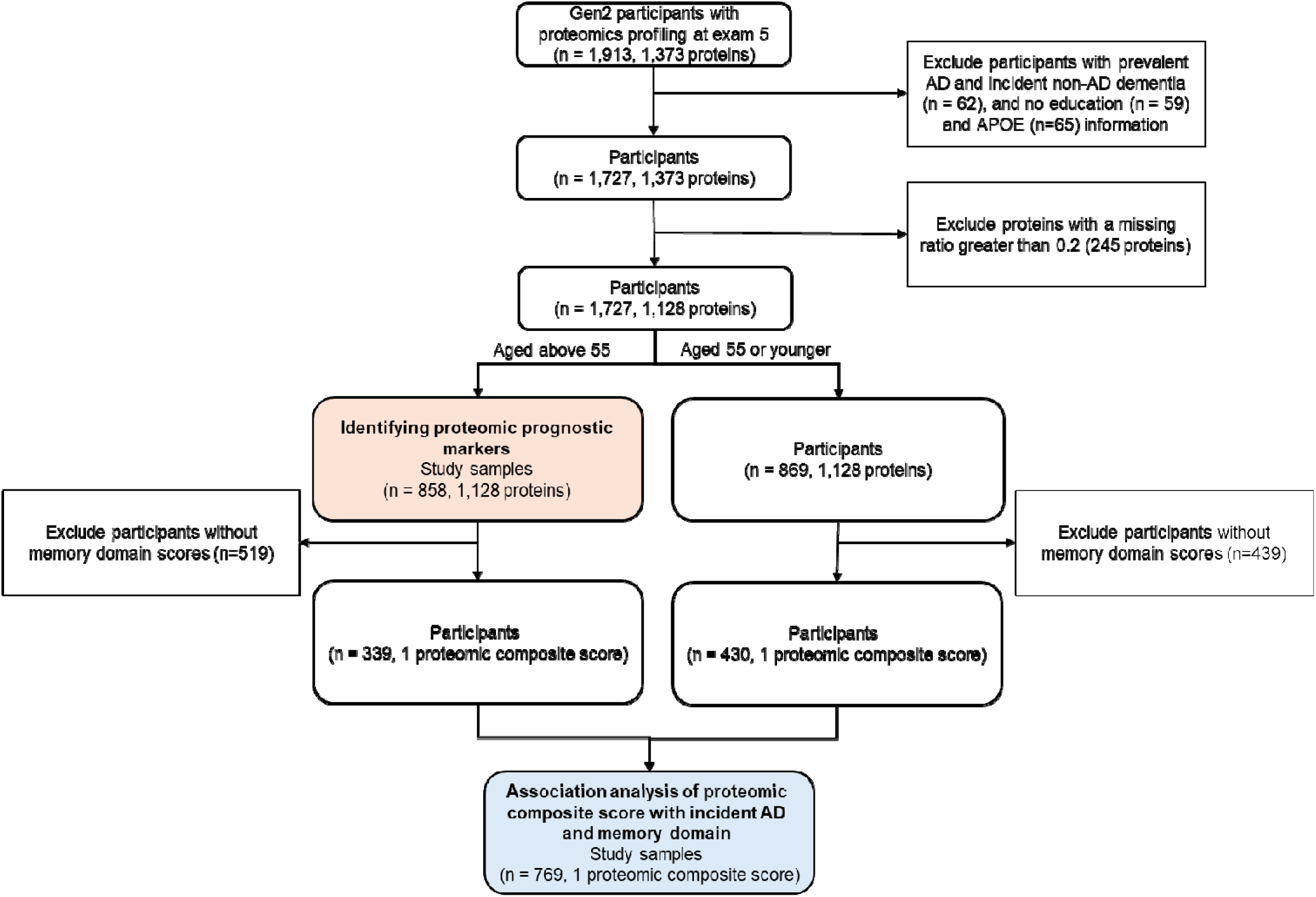
The sample selection flowchart of this study.

### Proteomics Profiling

Previous studies have detailed the methods of proteomics profiling[21, 22]. In brief, plasma was obtained from blood samples collected at clinical visits and preserved at -80°C[23]. The quantification of protein concentrations with these plasma samples was conducted using the SOMAscan platform[24]. This approach leverages single-stranded DNA aptamers to identify and bind to specific proteins. The efficacy of this technology has been validated through its application in cardiovascular disease research[21, 25]. The protein profiling analysis was conducted on the samples in two distinct batches with 821 and 1,092 participants, respectively. Across the two batches, a total of 1,373 proteins were examined. Logarithmic transformations were applied to the protein measurements, followed by inverse normal transformation to achieve normality. Linear models were used to obtain residuals by regressing the transformed proteins on Plate ID to minimize batch effect. Plate ID denotes the plate in the machine on which a given sample was run. Proteins (n=245) with more than 20% missing data were excluded (**Figure 1**). The residuals of protein markers were combined from two batches and a total of 1,128 proteins were used for subsequent analyses.

### Ascertainment of AD

A participant was identified with incident AD if they were cognitively intact at the time of proteomics profiling but diagnosed with AD during the follow-up. The methodology for monitoring and diagnosing AD within the FHS has been documented in prior publications[26, 27]. In brief, each participant diagnosed with AD was confirmed by a review panel comprising at least a neurologist and a neuropsychologist in FHS. This confirmation is based on available data from neurological and neuropsychological (NP) evaluations, FHS health examinations, clinical records, and discussions with relatives[28]. The criteria for AD diagnosis follow those established by the National Institute of Neurological and Communicative Disorders and Stroke and the Alzheimer’s Disease and Related Disorders Association (NINCDS–ADRDA)[29]. For the participants who developed AD, the follow-up duration was calculated from the baseline up to the earliest documented date of AD onset. For those who did not develop AD, the follow-up duration was terminated between the baseline and December 31 of 2022, the date of the last recorded follow-up, or the date of death, depending on which occurred first.

### Neuropsychological assessment

The administration of the neuropsychological tests in FHS has been detailed in prior studies [30, 31]. In brief, we obtained the z-scores of six Wechsler Memory Scale (WMS) scores, including WMS Logical Memory Immediate Recall, WMS Logical Memory Delayed Recall, WMS Visual Reproductions Immediate Recall, WMS Visual Reproductions Delayed Recall, WMS Paired Associates Immediate Recall, and WMS Paired Associates Delayed Recall [32, 33]. A z-score was calculated by subtract a score to its mean and divided by its standard deviation. The total memory domain score was the sum of the z-scores from the six WMS variables.

### Statistical Analyses

#### Descriptive statistics

This study conducted a comparative analysis of baseline characteristics between participants who developed AD during the follow-up period and those who did not. Continuous variables were compared using the t-test, and categorical variables were assessed using the Chi-square test to identify any significant differences between the two groups.

### Association Analysis and Survival Machine Learning for Identifying Proteomic Prognostic Markers

The association of each protein marker with incident AD was examined using Cox proportional hazard regression models. These models were adjusted for baseline demographic factors, including age, sex, education, and APOE ε4+ status. Proteins with a *P* value ≤0.2 were selected for further analysis with the machine learning methods.

This study evaluated the importance of proteins using two survival machine learning methods[34]: the LASSO-based Cox proportional hazard regression model (LASSO)[35] and the generalized boosted regression model (GBM) [36]. LASSO utilizes L1 regularization to induce sparsity, setting the coefficients of less important variables to zero, thus simplifying model complexity and preventing overfitting [37]. In contrast, GBM enhances model accuracy through an iterative process where each new model corrects errors from preceding ones, adeptly managing complex non-linear data patterns[38]. Both LASSO and GBM were adapted to accommodate censored data in analyzing time-to-event data. LASSO incorporates a Cox proportional hazards model, applying L1 penalties to enhance model selection and penalize less significant variables. GBM extends to survival scenarios by employing survival trees within its boosting framework. In determining protein importance, LASSO measures it by the magnitude of the coefficients, with larger values indicating a stronger impact on AD incidence[39]. GBM assesses protein importance by measuring its frequency in tree splitting and contribution to model performance, known as relative importance[40].

The survival LASSO and GBM models were developed using proteins with a *P* value ≤ 0.2. The proteins were initially ranked according to their importance. Beginning with the most significant protein marker, we sequentially built machine learning models, incorporating the next most important protein into each successive model. The mean Harrell’s c-index was calculated using a ten-fold cross-validation approach for each model iteration[41]. Ultimately, the final selected model for LASSO and GBM was determined by identifying the one with the fewest proteins among the top 5 models that achieved the highest mean Harrell’s c-index.

To minimize collinearity issue in regression model and bias in constructing a proteomic composite score, we calculated the pairwise Pearson correlation coefficients for each pair of proteins to eliminate highly correlated proteins identified by the machine learning models. For pairs where the correlation coefficient exceeded 0.3, only the protein with a more significant association with incident AD was retained as the proteomic prognostic marker.

### Construction of Proteomic Composite Score

To explore the cumulative impact of proteins on AD development, we constructed weighted composite scores using proteins that were previously identified by the machine learning models. The weights assigned to each protein were derived from their regression coefficients, obtained using a Cox proportional hazard model that adjusted for age, sex, education, and APOE ε4+ status. For proteins that achieved nominal significance (*P* < 0.05), the composite score was formulated as a linear combination of these weighted proteins.

### Association of Proteomic Composite Score with Incident AD and Memory Domain

The association between proteomic composite score and incident AD was examined by Cox proportional hazard regression model, adjusting for age, sex, education, and APOE ε4+ status. To further validate the early detection capabilities of the proteomic composite score for cognitive decline, we assessed its association with the memory domain score in a linear regression model, adjusting for age, sex, and education. We conducted the linear models in three groups of participants to investigate the association between the memory domain scores and composite scores, adjusting for age, sex, education, and APOE ε4+ status: 339 of the 858 participants with available memory domain scores (the older age group), a separate younger group of 430 independent participants (mean age 46, 56.7% women, the younger age group), and the pooled samples (n=769, the combined sample of 339 and 430 participants) (**Figure 1**).

### Assessment of AD Prediction Performance of Proteomic Composite Score

We compared the capacity of the proteomic composite score in predicting AD. The base model included age, sex, education, and APOE ε4+ status as predictors. A second model added proteins with *P*≤0.2 to the base model. The third model incorporated proteomic prognostic markers into the base model. The fourth model included the proteomic composite score into the base model. Both LASSO and GBM were utilized to evaluate these models. The performance of these models was evaluated using a 10-fold cross-validation. The time-dependent area under the receiver-operating characteristic curve (AUC) for each year during a follow-up period of 15 to 25 years was calculated to determine the model’s prediction performance [42].

## Results

### Baseline Demographics

To identify proteomic prognostic markers, this study included 858 FHS Offspring participants who were cognitively intact at baseline (mean age 63±5, 52.9% women, 34.0% college or above) (**Table 1**). During a mean follow-up of 20 years, 132 (15.4%) incident AD cases were identified.

**Table 1.** Baseline characteristics of study samples.

| Variable | Free from AD (N=726) | Incident AD (N=132) | Total (N=858) | *P* value<sup>*</sup> |
| --- | --- | --- | --- | --- |
| Age, years | 62.6 (5.4) | 65.4 (5.0) | 63.0 (5.4) | < 0.001 |
| Women, n(%) | 362 (49.9%) | 92 (69.7%) | 454 (52.9%) | < 0.001 |
| Education, n(%) |  |  |  | 0.187 |
| Less than high school | 66 (9.1%) | 15 (11.4%) | 81 (9.4%) |  |
| High school/some college | 404 (55.6%) | 81 (61.4%) | 485 (56.5%) |  |
| College or above | 256 (35.3%) | 36 (27.3%) | 292 (34.0%) |  |
| APOE ε4+, n(%) | 147 (20.2%) | 46 (34.8%) | 193 (22.5%) | < 0.001 |
| Follow-up, years | 20.1 (7.2) | 16.7 (6.1) | 19.6 (7.2) | < 0.001 |

Mean (standard deviation, SD) was presented for continuous variables and count (percentages) for categorical variables. \*Continuous variables were analyzed with the t-test, while categorical variables were examined using Chi-square test.

### The Association Between Proteins and Incident AD

Among the 1,128 proteins evaluated, 106 proteins were associated with incident AD with nominal significance after adjusting for age, sex, education, and APOE ε4+ status (*P*<0.05) (**Supplementary Table 1**). However, none of these proteins remain significant after false discovery rate correction. Among these, 73 proteins were positively associated with incident AD, with the most significant association identified in death-associated protein kinase 2 (DAPK2). Each SD higher level of DAPK2 was associated with a 44% higher risk of incident AD (95% CI: 1.19, 1.73; *P* = 8.27E-05). Conversely, 33 proteins showed negative associations with AD incidence, with the strongest association for hepatocyte growth factor receptors. Each SD increase in the plasma levels of hepatocyte growth factor receptor was associated with a 27% lower risk of incident AD (95% CI: 0.60, 0.89; *P* = 1.66E-03).

### Proteomic Prognostic Markers

For the 309 proteins associated with incident AD with a significance of *P*<0.2, we further evaluated their predictive capacity for AD using LASSO and GBM models. **Figure 2** presents the importance rankings of these proteins as determined by both models. Starting with the most significant protein, we incrementally added proteins to both the LASSO and GBM models to enhance AD prediction. **Figure 2** also illustrates the mean Harrell’s c-index from ten-fold cross-validation as additional proteins are incorporated into the models. The results indicated that the GBM model achieves its highest Harrell’s c-index, 0.750, when the top 19 proteins are included. The LASSO model achieved its optimal predictive performance with a Harrell’s c-index of 0.804 when 19 proteins were included. Five proteins, including GFRa-1, FCN1, Activated Protein C, Siglec-3, LIGHT, were identified by both the LASSO and GBM models. Therefore, a total of 33 proteins were identified by either LASSO or GBM. The univariate association of each of these 33 proteins with incident AD is shown in **Table 2**, with 6 of these proteins showing a negative association with AD.

**Figure 2.**
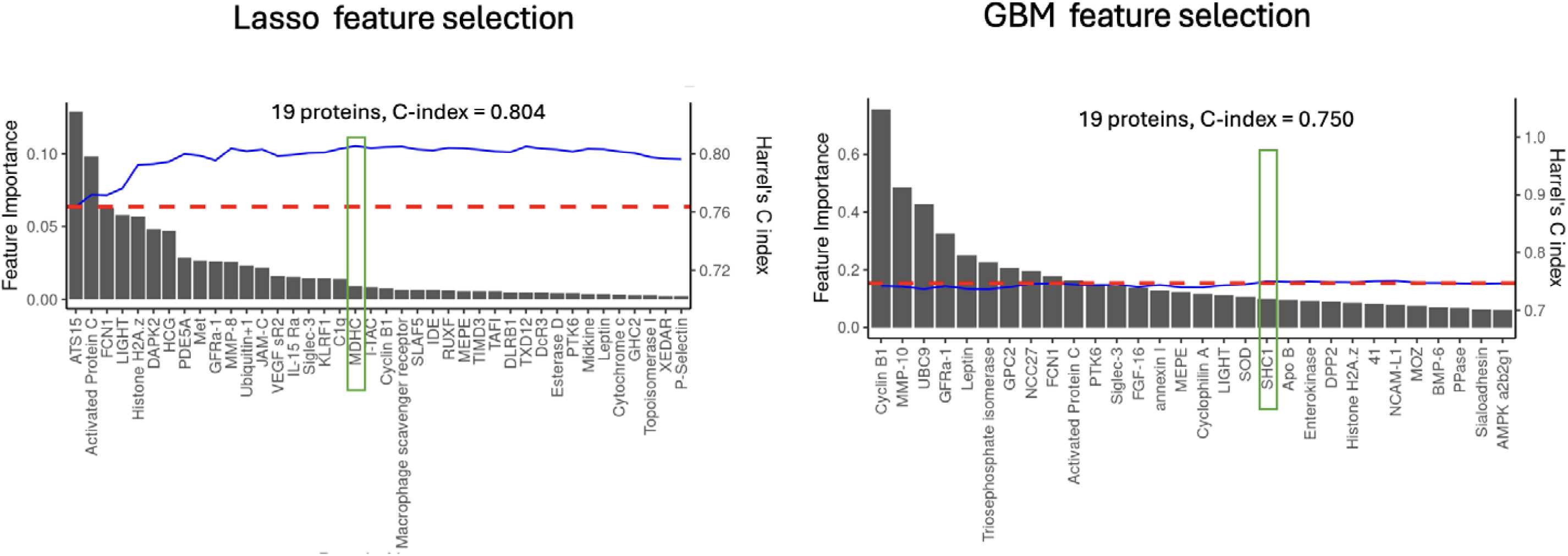
Protein selection by LASSO and GBM. The bar charts show the feature importance of each protein, while the blue line depict the Harrell’s c-index, illustrating how the model’s predictive performance improves with the sequential inclusion of proteins. Proteins ultimately selected are highlighted within a green box.

**Table 2.** Associations of 33 proteins selected by LASSO and GBM with incident AD.

| Protein name | Description | HR | 95% CI |  | P value |
| --- | --- | --- | --- | --- | --- |
| DAPK2 | Death-associated protein kinase 2 - 4355-13 (Q9UIK4) | 1.44 | 1.19 | 1.73 | 8.27E-05 |
| Activated Protein C | Activated Protein C - 3758-68 (P04070) | 1.36 | 1.16 | 1.59 | 1.02E-04 |
| ATS15 | A disintegrin and metalloproteinase with thrombospondin motifs 15 - 4533-76 (Q8TE58) | 1.43 | 1.18 | 1.75 | 2.76E-04 |
| Histone H2A.z | Histone H2A.z - 4163-5 (P0C0S5) | 1.4 | 1.16 | 1.69 | 3.26E-04 |
| FCN1 | Ficolin-1 - 3613-62 (O00602) | 1.38 | 1.15 | 1.66 | 4.84E-04 |
| Met | Hepatocyte growth factor receptor - 2837-3 (P08581) | 0.73 | 0.6 | 0.89 | 1.66E-03 |
| LIGHT | Tumor necrosis factor ligand superfamily member 14 - 5355-69 (O43557) | 1.32 | 1.1 | 1.58 | 1.81E-03 |
| Ubiquitin+1 | Ubiquitin+1, truncated mutation for UbB - 2846-24 (P62979) | 1.3 | 1.09 | 1.55 | 3.51E-03 |
| MDHC | Malate dehydrogenase, cytoplasmic - 3853-56 (P40925) | 1.32 | 1.09 | 1.61 | 4.59E-03 |
| C1q | Complement C1q subcomponent - 2753-2 (P02745 P02746 P02747) | 0.77 | 0.63 | 0.94 | 9.99E-03 |
| Cyclin B1 | G2/mitotic-specific cyclin-B1 - 5347-59 (P14635) | 1.24 | 1.05 | 1.47 | 1.11E-02 |
| PTK6 | Protein-tyrosine kinase 6 - 3832-51 (Q13882) | 0.79 | 0.66 | 0.95 | 1.11E-02 |
| JAM-C | Junctional adhesion molecule C - 2998-53 (Q9BX67) | 1.27 | 1.05 | 1.54 | 1.15E-02 |
| GFRa-1 | GDNF family receptor alpha-1 - 3314-74 (P56159) | 1.28 | 1.05 | 1.55 | 1.18E-02 |
| VEGF sR2 | Vascular endothelial growth factor receptor 2 - 3651-50 (P35968) | 0.78 | 0.64 | 0.95 | 1.38E-02 |
| SOD | Superoxide dismutase [Cu-Zn] - 2794-60 (P00441) | 1.26 | 1.04 | 1.52 | 1.44E-02 |
| IL-15 Ra | Interleukin-15 receptor subunit alpha - 3445-53 (Q13261) | 1.27 | 1.04 | 1.56 | 1.81E-02 |
| PDE5A | cGMP-specific 3,5-cyclic phosphodiesterase - 5256-86 (O76074) | 1.24 | 1.02 | 1.49 | 2.37E-02 |
| HCG | Human Chorionic Gonadotropin - 4914-10 (P01215,P01233) | 1.33 | 1.03 | 1.72 | 2.74E-02 |
| Leptin | Leptin - 2575-5 (P41159) | 1.27 | 1.02 | 1.58 | 2.82E-02 |
| MMP-8 | Neutrophil collagenase - 2954-56 (P22894) | 1.21 | 1 | 1.47 | 4.27E-02 |
| NCC27 | Chloride intracellular channel protein 1 - 5013-2 (O00299) | 1.19 | 1 | 1.43 | 4.69E-02 |
| KLRF1 | Killer cell lectin-like receptor subfamily F member 1 - 5098-79 (Q9NZS2) | 1.2 | 1 | 1.44 | 5.06E-02 |
| MMP-10 | Stromelysin-2 - 3743-1 (P09238) | 1.18 | 0.99 | 1.41 | 5.25E-02 |
| Triosephosphate isomerase | Triosephosphate isomerase - 4309-59 (P60174) | 1.18 | 0.99 | 1.42 | 6.40E-02 |
| UBC9 | SUMO-conjugating enzyme UBC9 - 2877-3 (P63279) | 1.19 | 0.99 | 1.44 | 6.48E-02 |
| annexin I | Annexin A1 - 4960-72 (P04083) | 1.18 | 0.98 | 1.43 | 8.11E-02 |
| Siglec-3 | Myeloid cell surface antigen CD33 - 3166-92 (P20138) | 1.16 | 0.97 | 1.38 | 8.83E-02 |
| Cyclophilin A | Peptidyl-prolyl cis-trans isomerase A - 3844-2 (P62937) | 1.18 | 0.97 | 1.44 | 1.00E-01 |
| MEPE | Matrix extracellular phosphoglycoprotein - 3209-69 (Q9NQ76) | 0.85 | 0.71 | 1.03 | 1.01E-01 |
| FGF-16 | Fibroblast growth factor 16 - 4393-3 (O43320) | 1.15 | 0.95 | 1.4 | 1.42E-01 |
| SHC1 | SHC-transforming protein 1 - 5272-55 (P29353) | 1.15 | 0.95 | 1.41 | 1.49E-01 |
| GPC2 | Glypican-2 - 3315-15 (Q8N158) | 0.88 | 0.74 | 1.05 | 1.53E-01 |

Note: The association of incident AD with each protein was examined by Cox proportional hazards model adjusting for baseline age, sex, education, and APOE ε4+ status.

**Figure 3** displays a heatmap of the Pearson correlation coefficients for the 33 proteins. Among these, 15 pairs of proteins exhibited correlation coefficients greater than 0.3. Consequently, we eliminated the less significant proteins associated with incident AD from these 15 pairs. After this filtering process, 18 proteins were retained for further analysis.

**Figure 3.**
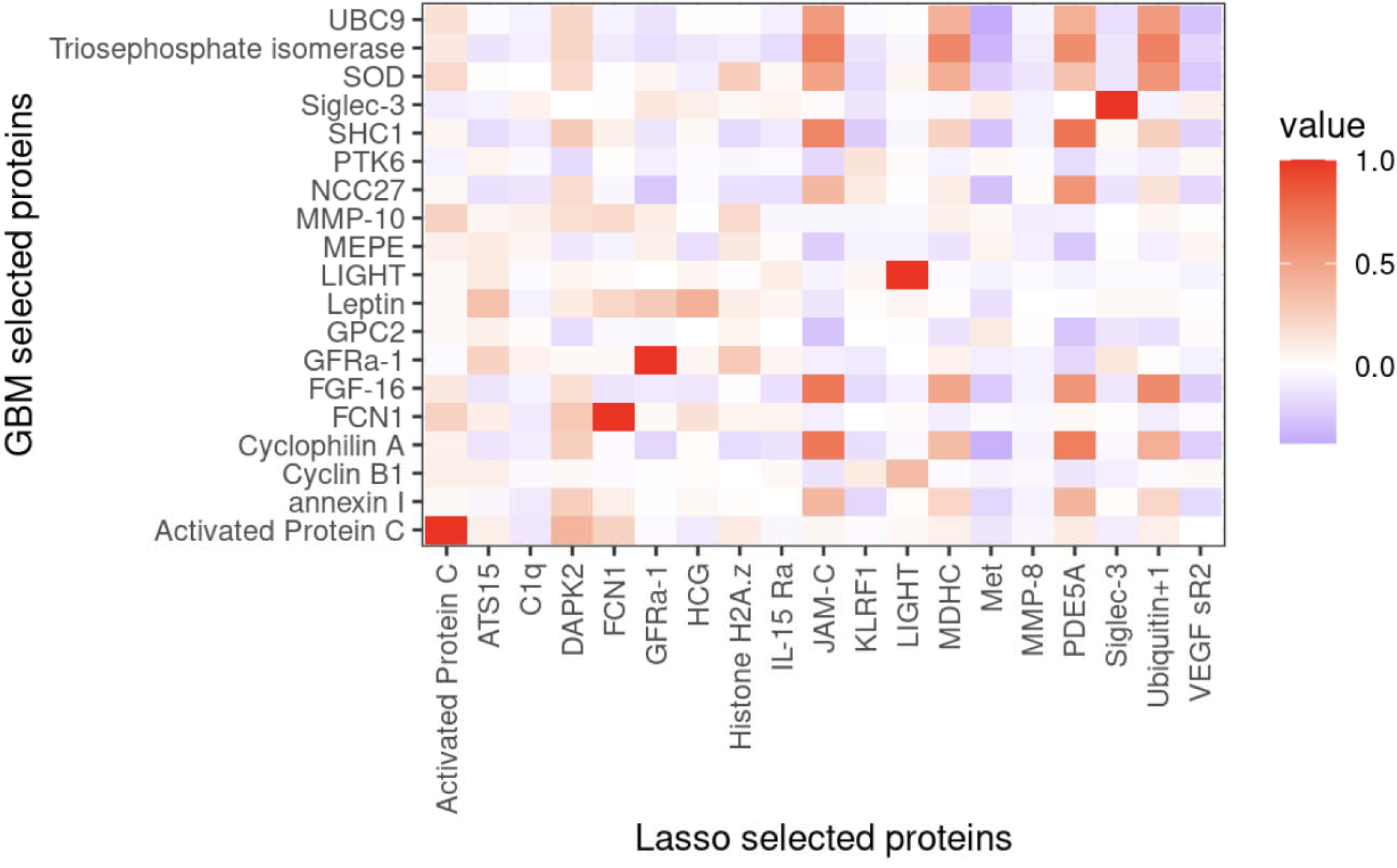
The correlation of proteins selected by LASSO and GBM. The color gradient represents the correlation levels, where darker red indicates a higher positive correlation and lighter pink signifies a higher negative correlation between the protein pairs selected by the two methods.

### Association Analysis of Proteomic Composite Score with the Memory Domain Score

A multivariable Cox regression model assessed the associations between 18 proteins and incident AD, adjusting for age, sex, education, and APOE ε4+ status as covariates. In this model, 9 proteins demonstrated significance with a *P* value less than 0.05. Consequently, these 9 proteins were used to construct a composite score (**Figure 4**). In association analyses, the composite score was positively associated with the incidence of AD. Each unit increase in the composite score was associated with a 2.3 times higher risk of developing AD (HR = 2.33; 95% CI: 1.85, 2.79; *P* = 5.8E-15). To test if the proteomic composite score was predictive to memory score, we conducted a linear regression and found that the proteomic composite score was negatively associated with the memory domain score in the 339 (of the 858) participants with available memory scores (beta= -0.061, SE = 0.030, *P* = 0.046), adjusting for age, sex, education, and APOE ε4+ status (**Figure 5**). This negative association was consistent in the independent 430 participants aged 55 and below (beta = -0.060, SE = 0.025, *P* = 0.018), in the pooled 769 samples (beta = -0.058, SE = 0.019, *P* = 0.003), adjusting for age, sex, education, and APOE ε4+ status (**Figure 5**).

**Figure 4.**
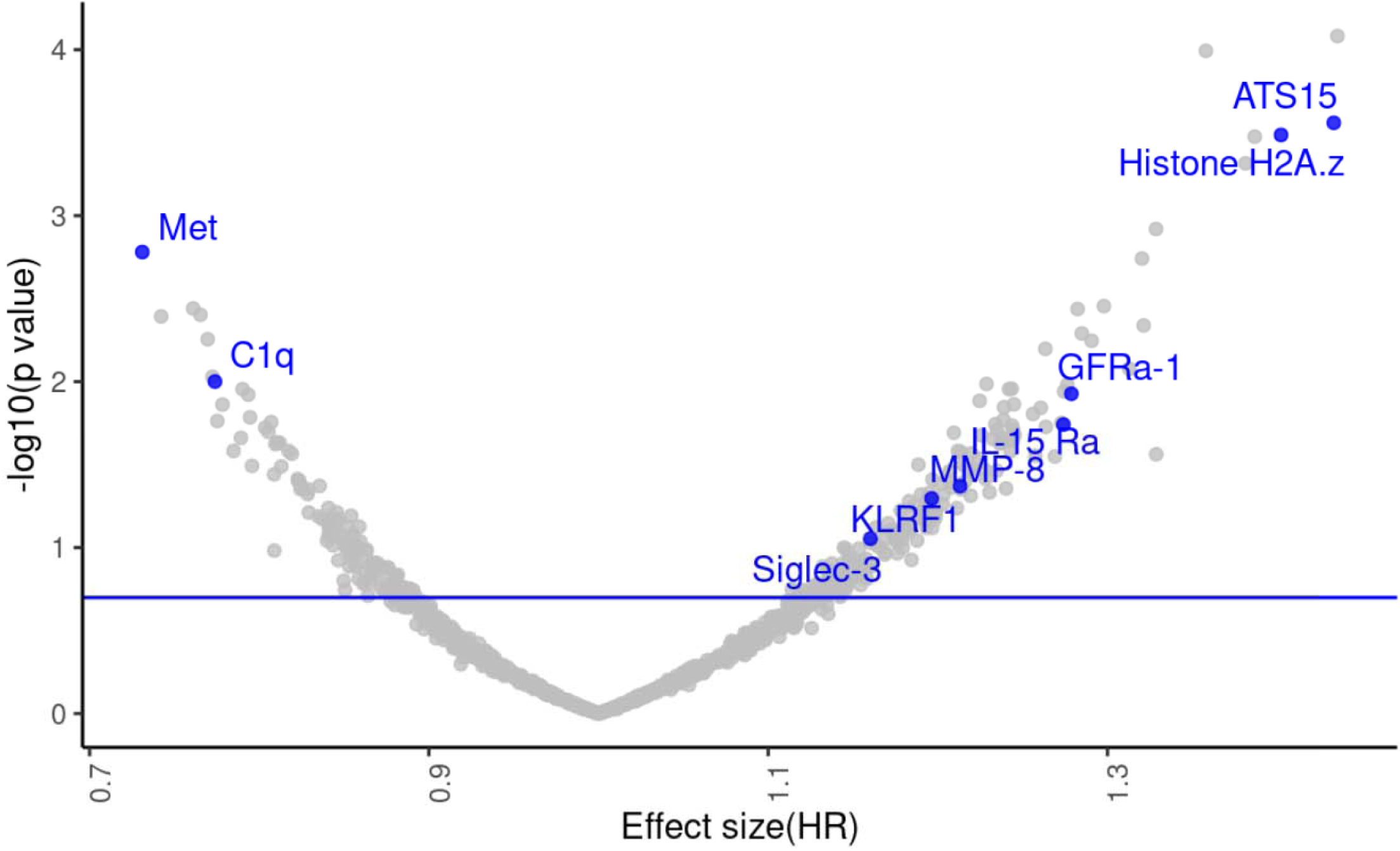
Volcano plots illustrate the HR on the x-axis and -log10(*P* value) on the y-axis, showing the association of proteins with incident AD. Proteins located above the horizontal blue line indicate the significance (*P* < 0.2) in their association with incident AD. The 9 selected proteomic prognostic markers were highlighted with blue dots.

**Figure 5.**
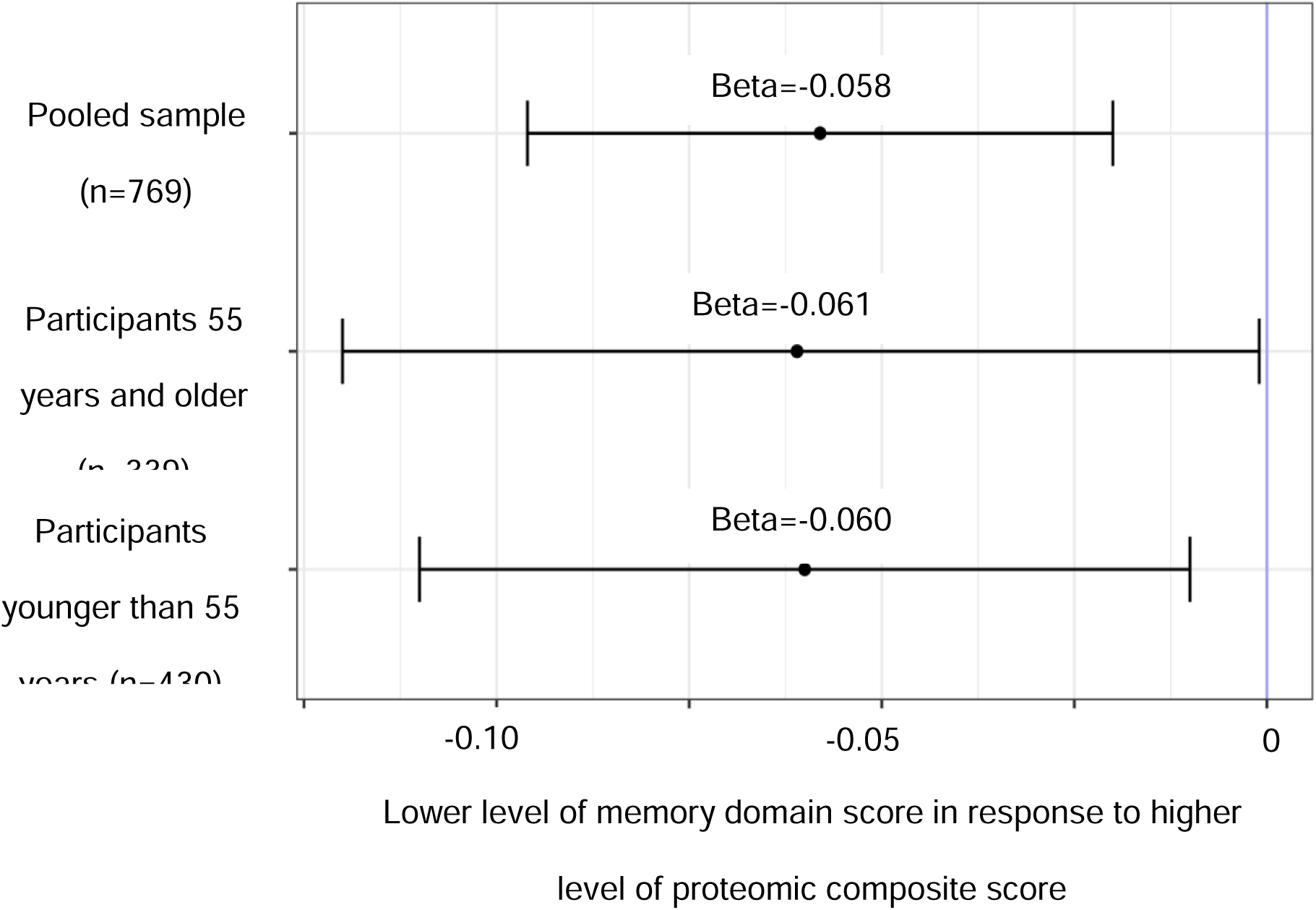
Forest plot showing the association of memory domain score with proteomic composite score. The association was examined by linear regression model adjusting for baseline age, sex, education, APOE ε4+ status. SE: standard error.

### Performance Comparison of AD Predictive Models Across Varying Follow-Up Periods

We evaluated the prediction performance of incident AD using different models over a follow-up time ranging from 15 to 25 years (**Figure 6**). In the GBM analysis, Model 4, which integrates age, sex, education, and APOE ε4+ status with a proteomic composite score, consistently outperforms the other models, maintaining AUC values above 0.797 throughout the period. Model 1, which includes only age, sex, education, and APOE ε4+ status, ranks as the second-best model, achieving its optimal AUC of 0.800 at the 21-year follow-up. Similar patterns are observed with the LASSO model, where Model 4 also consistently achieves the highest AUC. Additionally, Models 2 and 3 which incorporated proteins, generally remain above the performance of Model 1 (optimal AUC 0.826 at 15-year follow-up). For Model 4 fitting with LASSO, the predictive performance for AD starts with an AUC of 0.83 at the 15-year follow-up, peaks at 0.84 at the 22-year follow-up, and then generally shows a declining trend in predictive performance as the follow-up period extends (mean AUC: 0.79).

**Figure 6.**
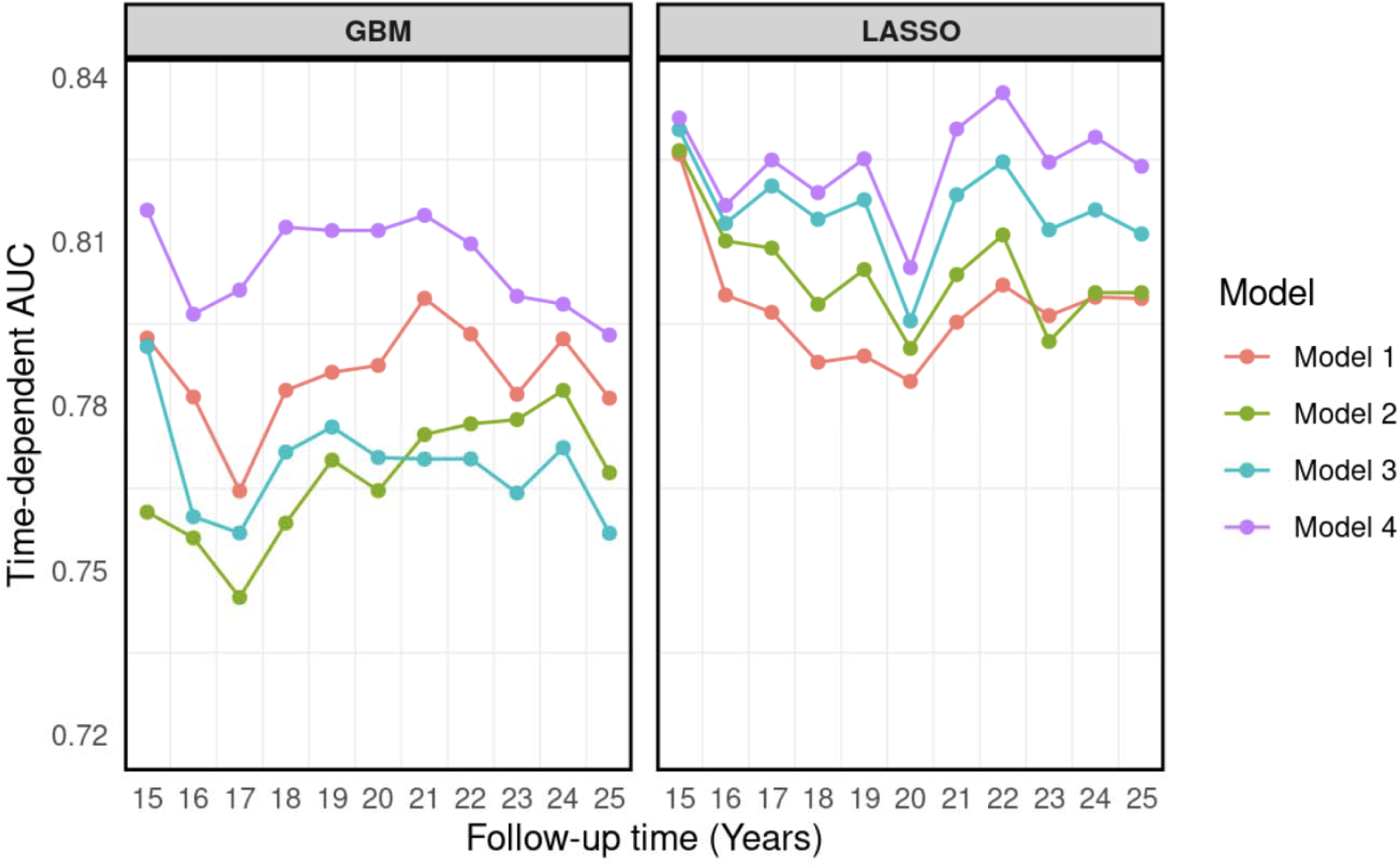
Time-dependent AUC estimates of different models at each year of follow-up between 15 years and 25 years. Model 1: age, sex, education, and APOE ε4+ status; Model 2: age, sex, education, APOE ε4+ status, and 309 proteins; Model 3: age, sex, education, APOE ε4+ status, and 9 proteomic prognostic markers; Model 4: age, sex, education, APOE ε4+ status, and proteomic composite score. The horizontal axis represents follow-up time in years and the vertical axis represents the estimated area under the ROC curve for survival at the time of interest.

## Discussion

This study identified a set of proteomic prognostic markers for AD by utilizing association analysis and survival machine learning models. A proteomic composite score was developed from 9 selected proteins, which, when integrated with clinical risk factors, significantly enhanced the performance of models for AD prediction across various follow-up periods, reaching a peak AUC of 0.84 at the 22-year follow-up. To our knowledge, this is the first study to employ a survival machine learning-based approach to identify proteomic prognostic markers for AD, consistently achieving high predictive results from 15 to 25 years.

Identifying prognostic markers capable of predicting AD risk across various follow-up periods is crucial for both understanding the disease’s progression and selecting participants for clinical trials focused on preventive therapies. Proteins with their ability to reflect biological processes and responses to treatments makes them particularly valuable in tracking the gradual development of AD and assessing intervention efficacy over time. In this study, 106 out of 1,128 proteins showed nominal significance (*P* < 0.05) in association with incident AD after adjusting for baseline demographics. The most significant association was observed between DAPK2 and the risk of AD. DAPK2 shares a high degree of homology with DAPK1, particularly in their catalytic domains[43]. DAPK1 is widely expressed throughout the central nervous system and its dysregulation has been linked to neurological disorders, including AD[44]. Notably, activated protein C emerged as significantly associated with an increased risk of AD, despite its known neuroprotective effects, including anti-inflammatory properties and roles in promoting blood flow and preventing brain cell death [45]. This indicates that elevated levels of activated protein C might be a compensatory response to early pathophysiological changes in AD, where the body attempts to counteract damage but may inadvertently exacerbate other underlying mechanisms that promote AD progression. Another possible reason could be the presence of other confounding factors affecting its association with AD. Conversely, hepatocyte growth factor receptor showed the most significant negative association with AD incidence, aligning with findings from other research [46]. Activation of hepatocyte growth factor facilitates stem cell differentiation and neurogenesis, and offers protection against damage in various cells, including neurons[47]. These findings not only contribute to our understanding of the biological pathways involved in AD but also underscore the complexity of the disease’s pathophysiology, suggesting that both inflammatory processes and regenerative mechanisms may play crucial roles.

The use of survival machine learning models to further refine the selection of proteomic markers based on their predictive relevance represents an advanced approach to handling high-dimensional data and incorporating the impact of time. Traditional methods like stepwise selection, which assume independence among predictors, may be biased given correlation among proteins. In contrast, LASSO addresses this issue by employing L1 optimization, which minimizes the total sum of coefficients, selecting one protein from a highly correlated pair. However, due to the high degree of correlation (or collinearity) among proteins, LASSO may randomly select one protein from correlated pairs, resulting in variability with each run. To minimize the issue, we implemented cross-validation to average the feature importance of each protein, providing a more stable and reliable approach to manage collinearity. Additionally, we employed the GBM, a tree-based ensemble method that uses a boosting framework to train models iteratively. Unlike methods that rely on single-variable fitting, GBM utilizes a subset of variables for each tree, enhancing its capacity to integrate and analyze proteins collectively. This method effectively captures the complex interdependencies among proteins and enhances the model’s predictive performance for AD by leveraging multiple variables simultaneously. The ability of these models to achieve high Harrell’s c-index with a relatively small subset of proteins suggests that machine learning techniques can effectively extract key information about the relationship between proteins and AD risk. Notably, both models consistently identified a core set of five proteins, including GFRa-1, FCN1, Activated Protein C, Siglec-3, LIGHT, that share predictive relevance for AD, providing a strong validation for these findings and underscoring the robustness of these methods in identifying critical markers. This alignment with prior research further reinforces the significance of these proteins in AD. For example, GFRa-1 is implicated in neural cell survival and repair mechanisms, highlighting its potential role in AD [48]. FCN1 (Ficolin-1) has been shown to be differentially expressed in AD cases[49]. Siglec-3, expressed in myeloid cells, plays a role in the immune response of neurodegenerative diseases [50]. Tumor necrosis factor has been shown to be associated with AD[51].

Researchers have been investigating the potential of plasma proteomic profiles to predict incident AD[13, 52]. In this study, we developed a proteomic composite score from ten proteomic prognostic markers and evaluated its association with AD incidence. This score positively associated with AD risk, suggesting it could be a crucial part of a broader diagnostic framework to identify individuals at increased risk of AD before symptoms appear. Previous studies have focused on predicting AD incidence at a single future time point[13, 52]. In contrast, our study extends this approach by assessing the composite score’s ability to predict AD incidence over multiple follow-up periods. The enhanced predictive performance, evidenced by increased time-dependent AUC values when integrating proteomic markers with baseline demographics, is particularly noteworthy. Additionally, we calculated the weighted composite scores for individuals under 55 years of age with regression coefficients obtained in the older age group and analyzed its association with memory domain scores. The consistent significant negative association with the general population indicates that the composite score is effective at detecting early changes in cognitive function. The validation of the proteomic composite score is warranted in future studies with larger sample sizes of both older and younger age groups.

We recognize several limitations in our study. Our participants consisted solely of non-Hispanic whites, highlighting the need for future research in diverse ethnic and racial groups. Moreover, despite the robust predictive performance of our selected markers across multiple follow-up times, potential confounders could still influence the association between the proteins and incident AD. Future studies should consider incorporating additional factors to address these potential confounders. This expansion is crucial for enhancing the generalizability of our findings and facilitating external validation. Moreover, similar to other high-throughput “omics” studies, batch effects could affect the reproducibility of our findings. Furthermore, although our proteomics platform is one of the most comprehensive available, it is limited to detecting only the proteins that are incorporated into the platform. Future studies should be conducted to validate our findings in larger cohorts. This study has several advantages. First, the integration of survival machine learning with association analysis enables the selection of markers that can accommodate the complex interactions within high-dimensional proteomic data and include time-to-event information. Second, this study developed protein composite scores that are highly interpretable and easy to use. Further, we validated these composite scores by examining their association with memory domain scores in a separate younger group. This demonstrated the effectiveness of the scores in detecting early cognitive changes. We utilized time-dependent AUC to assess the predictive power of protein composite score for AD incidence risk across various follow-up periods. This method allowed us to track how the prognostic abilities of these markers evolved over time, offering a comprehensive assessment of their long-term efficacy in predicting AD progression.

In summary, this study significantly advances the identification and application of AD proteomic prognostic markers through survival machine learning methods. It demonstrated the proteomic composite score’s ability to predict AD risk consistently across multiple follow-up periods. Further studies involving external validation are essential to ensure the generalizability of these findings.

## Data Availability

The data could be requested through an application to the FHS.

https://www.framinghamheartstudy.org/fhs-for-researchers

## Declarations

### Ethics approval and consent to participate

All participants provided their written consent for genetic studies. The study protocol received approval from the Institutional Review Boards at Boston University Medical Center, Massachusetts General Hospital, and Beth Israel Deaconess Medical Center. The study adhered strictly to regulations and guidelines to ensure compliance.

### Availability of data and materials

The data could be requested through an application to the FHS (https://www.framinghamheartstudy.org/fhs-for-researchers).

### Authors’ contributions

CL designed the study. YL analyzed the data and trained the model. HD made significant contributions to writing the manuscript and interpreting the results. TA, RA, and MD reviewed and edited the manuscript. All authors read and approved the final manuscript.

## Acknowledgments

We would like to thank the participants of the Framingham Heart Study for their commitment. This research could not have been conducted without their involvement.

## References

1. Hroudová, J., N. Singh, and Z. Fišar, Mitochondrial dysfunctions in neurodegenerative diseases: relevance to Alzheimer’s disease. BioMed research international, 2014. 2014.

2. Giebel, C.M., C. Sutcliffe, and D. Challis, Activities of daily living and quality of life across different stages of dementia: a UK study. Aging & mental health, 2015. 19(1): p. 63–71.

3. Jones, R.W., et al., Dependence in Alzheimer’s disease and service use costs, quality of life, and caregiver burden: the DADE study. Alzheimer’s & Dementia, 2015. 11(3): p. 280–290.

4. Nichols, E., et al., Estimation of the global prevalence of dementia in 2019 and forecasted prevalence in 2050: an analysis for the Global Burden of Disease Study 2019. The Lancet Public Health, 2022. 7(2): p. e105–e125.

5. Abdelnour, C., et al., Perspectives and challenges in patient stratification in Alzheimer’s disease. Alzheimer’s research & therapy, 2022. 14(1): p. 112.

6. Jack, C.R., et al., Hypothetical model of dynamic biomarkers of the Alzheimer’s pathological cascade. The Lancet Neurology, 2010. 9(1): p. 119–128.

7. Rockwood, K., Biomarkers to measure treatment effects in Alzheimer’s disease: what should we look for? International Journal of Alzheimer’s Disease, 2011. 2011.

8. Bangma, J., et al., Understanding the dynamics of physiological changes, protein expression, and PFAS in wildlife. Environment international, 2022. 159: p. 107037.

9. Walker, K.A., et al., Large-scale plasma proteomic analysis identifies proteins and pathways associated with dementia risk. Nature Aging, 2021. 1(5): p. 473–489.

10. Nazeri, A., et al., Imaging proteomics for diagnosis, monitoring and prediction of Alzheimer’s disease. Neuroimage, 2014. 102: p. 657–665.

11. Sattlecker, M., et al., Alzheimer’s disease biomarker discovery using SOMAscan multiplexed protein technology. Alzheimer’s & Dementia, 2014. 10(6): p. 724–734.

12. Bai, B., et al., Proteomic landscape of Alzheimer’s Disease: novel insights into pathogenesis and biomarker discovery. Molecular neurodegeneration, 2021. 16(1): p. 55.

13. Hye, A., et al., Proteome-based plasma biomarkers for Alzheimer’s disease. Brain, 2006. 129(11): p. 3042–3050.

14. Shi, L., et al., Discovery and validation of plasma proteomic biomarkers relating to brain amyloid burden by SOMAscan assay. Alzheimer’s & Dementia, 2019. 15(11): p. 1478–1488.

15. Wang, P., Y. Li, and C.K. Reddy, Machine learning for survival analysis: A survey. ACM Computing Surveys (CSUR), 2019. 51(6): p. 1–36.

16. Hu, C. and J.A. Steingrimsson, Personalized risk prediction in clinical oncology research: applications and practical issues using survival trees and random forests. Journal of biopharmaceutical statistics, 2018. 28(2): p. 333–349.

17. Dawber, T.R., G.F. Meadors, and F.E. Moore Jr, Epidemiological approaches to heart disease: the Framingham Study. American Journal of Public Health and the Nations Health, 1951. 41(3): p. 279–286.

18. Kannel, W.B., et al., An investigation of coronary heart disease in families: the Framingham Offspring Study. American journal of epidemiology, 1979. 110(3): p. 281–290.

19. Yeon, S.B., et al., Impact of age, sex, and indexation method on MR left ventricular reference values in the Framingham Heart Study offspring cohort. Journal of Magnetic Resonance Imaging, 2015. 41(4): p. 1038–1045.

20. Johnson, J.M. and T.M. Khoshgoftaar, Survey on deep learning with class imbalance. Journal of Big Data, 2019. 6(1): p. 1–54.

21. Ngo, D., et al., Aptamer-based proteomic profiling reveals novel candidate biomarkers and pathways in cardiovascular disease. Circulation, 2016. 134(4): p. 270–285.

22. Hathout, Y., et al., Large-scale serum protein biomarker discovery in Duchenne muscular dystrophy. Proceedings of the National Academy of Sciences, 2015. 112(23): p. 7153–7158.

23. Benson, M.D., et al., Genetic architecture of the cardiovascular risk proteome. Circulation, 2018. 137(11): p. 1158–1172.

24. Gold, L., et al., Aptamer-based multiplexed proteomic technology for biomarker discovery. Nature Precedings, 2010: p. 1–1.

25. Ganz, P., et al., Development and validation of a protein-based risk score for cardiovascular outcomes among patients with stable coronary heart disease. Jama, 2016. 315(23): p. 2532–2541.

26. Bachman, D., et al., Prevalence of dementia and probable senile dementia of the Alzheimer type in the Framingham Study. Neurology, 1992. 42(1): p. 115–115.

27. Satizabal, C.L., et al., Incidence of dementia over three decades in the Framingham Heart Study. New England Journal of Medicine, 2016. 374(6): p. 523–532.

28. Au, R., et al., The Framingham Brain Donation Program: neuropathology along the cognitive continuum. Current Alzheimer Research, 2012. 9(6): p. 673–686.

29. McKhann, G.M., et al., The diagnosis of dementia due to Alzheimer’s disease: Recommendations from the National Institute on Aging-Alzheimer’s Association workgroups on diagnostic guidelines for Alzheimer’s disease. Alzheimer’s & dementia, 2011. 7(3): p. 263–269.

30. Au, R., R.J. Piers, and S. Devine, How technology is reshaping cognitive assessment: Lessons from the Framingham Heart Study. Neuropsychology, 2017. 31(8): p. 846–861.

31. Ferretti, M.T., et al., Maximizing utility of neuropsychological measures in sex-specific predictive models of incident Alzheimer’s disease in the Framingham Heart Study. Alzheimer’s & Dementia, 2023.

32. Ding, H., et al., Sex-specific blood biomarkers linked to memory changes in middle-aged adults: The Framingham Heart Study. Alzheimer’s & Dementia: Diagnosis, Assessment & Disease Monitoring, 2024. 16(1): p. e12569.

33. Wechsler, D., & Stone C. P, Wechsler Memory Scale (WMS). New York: The Psychological Corporation, 1948.

34. Raphael Sonabend, P.S., Sebastian Fischer, mlr3extralearners: Extra Learners For mlr3. 2024.

35. Simon, N., et al., Regularization paths for Cox’s proportional hazards model via coordinate descent. Journal of statistical software, 2011. 39(5): p. 1.

36. Ridgeway, G., Generalized Boosted Models: A guide to the gbm package. Update, 2007. 1(1): p. 2007.

37. Fonti, V. and E. Belitser, Feature selection using lasso. VU Amsterdam research paper in business analytics, 2017. 30: p. 1–25.

38. Zhang, Y. and A. Haghani, A gradient boosting method to improve travel time prediction. Transportation Research Part C: Emerging Technologies, 2015. 58: p. 308–324.

39. Muthukrishnan, R. and R. Rohini. LASSO: A feature selection technique in predictive modeling for machine learning. in 2016 IEEE international conference on advances in computer applications (ICACA). 2016. Ieee.

40. Friedman, J.H., Greedy function approximation: a gradient boosting machine. Annals of statistics, 2001: p. 1189–1232.

41. Harrell Jr, F.E., K.L. Lee, and D.B. Mark, Multivariable prognostic models: issues in developing models, evaluating assumptions and adequacy, and measuring and reducing errors. Statistics in medicine, 1996. 15(4): p. 361–387.

42. Chambless, L.E. and G. Diao, Estimation of time-dependent area under the ROC curve for long-term risk prediction. Statistics in medicine, 2006. 25(20): p. 3474–3486.

43. Bialik, S. and A. Kimchi, The death-associated protein kinases: structure, function, and beyond. Annu. Rev. Biochem., 2006. 75(1): p. 189–210.

44. Zhang, T., B.M. Kim, and T.H. Lee, Death-associated protein kinase 1 as a therapeutic target for Alzheimer’s disease. Translational neurodegeneration, 2024. 13(1): p. 4.

45. Shibata, M., et al., Anti-inflammatory, antithrombotic, and neuroprotective effects of activated protein C in a murine model of focal ischemic stroke. Circulation, 2001. 103(13): p. 1799–1805.

46. Wei, J., et al., Reduced HGF/MET signaling may contribute to the synaptic pathology in an Alzheimer’s disease mouse model. Frontiers in Aging Neuroscience, 2022. 14: p. 954266.

47. Wright, J.W. and J.W. Harding, The brain hepatocyte growth Factor/c-Met receptor system: A new target for the treatment of Alzheimer’s disease. Journal of Alzheimer’s Disease, 2015. 45(4): p. 985–1000.

48. Pöyhönen, S., et al., Effects of neurotrophic factors in glial cells in the central nervous system: expression and properties in neurodegeneration and injury. Frontiers in physiology, 2019. 10: p. 422442.

49. Zhang, X., et al., Identification of serum biomarkers in patients with Alzheimer’s disease by 2D-DIGE proteomics. Gerontology, 2022. 68(6): p. 686–698.

50. Siew, J.J., et al., Roles of Siglecs in neurodegenerative diseases. Molecular Aspects of Medicine, 2023. 90: p. 101141.

51. De Sousa Rodrigues, M.E., et al., Targeting soluble tumor necrosis factor as a potential intervention to lower risk for late-onset Alzheimer’s disease associated with obesity, metabolic syndrome, and type 2 diabetes. Alzheimer’s research & therapy, 2020. 12: p. 1–16.

52. Guo, Y., et al., Plasma proteomic profiles predict future dementia in healthy adults. Nature Aging, 2024: p. 1–14.

